# Prevalence and Factors Associated with Malnutrition Among Adult Cancer Patients Undergoing Treatment at KCMC Hospital, Kilimanjaro: A Facility-Based Cross-Sectional Study

**DOI:** 10.1101/2025.07.14.25331507

**Authors:** Ephraim E. Ng’weshemi, Furaha Serventi, Esther L. Majaliwa, Mary Mosha

## Abstract

**Background:** Globally, malnutrition affects 20%–70% of cancer patients and contributes to 10%–20% of cancer-related deaths, worsening treatment outcomes and survival. Despite its high prevalence, particularly in low-resource settings like Tanzania, research on adult cancer patients remains limited. This study aimed to determine the prevalence and factors associated with malnutrition among adult cancer patients undergoing treatment at Kilimanjaro Christian Medical Centre (KCMC) in Tanzania.

**Method:** A hospital-based cross-sectional study was conducted among 323 adult cancer patients (≥18 years) at KCMC Oncology Clinic from June to July 2024. Anthropometric measurements followed WHO standards, and data were collected using a modified Patient-Generated Subjective Global Assessment (PG-SGA) tool. Data analysis was performed using STATA version 15.0, whereby a log-binomial model was used to assess associations between predictor variables and malnutrition, with statistical significance set at p < 0.05.

**Results:** Out of 323 adult cancer patients, the average age was 59.6 years, with the majority being aged 60 years or older (53.9%). Females comprised 169 (52.3%), and males made up 154 (47.7%) of the sample. In terms of education, 170 (52.6%) had primary education, while 139 (43.3%) had higher education. Most patients were married, 218 (67.7%), and resided in rural areas, 179 (55.4%). Regarding employment status, 191 (59.1%) were unemployed, and 188 (58.2%) had health insurance. Among the 323 patients, 72.5% (234) were malnourished, including 72.8% (123) of females and 72.1% (111) of males. However, there was no significant association between the predictor variables and malnutrition.

**Conclusion:** This study found a high prevalence of malnutrition (72.5%) among adult cancer patients undergoing treatment at KCMC, with no significant associations observed between the predictor variables and malnutrition. This highlights the urgent need for targeted interventions to address malnutrition in cancer patients, particularly in low-resource settings like Tanzania. However, Further research is needed to better understand the impact of malnutrition on cancer treatment outcomes.

## Background

Cancer is a large group of diseases that can start in almost any organ or tissue of the body when abnormal cells grow uncontrollably, go beyond their usual boundaries to invade adjoining parts of the body, and/or spread to other organs[1]. It is the second leading cause of death globally, accounting for an estimated 9.6 million deaths, or 1 in 6 deaths[2]. The World Health Organization projects that the occurrence of cancer will increase exponentially by the year 2030, with the annual number of new cases rising from 14.1 million in 2012 to 21.6 million in 2030 and deaths due to cancer rising from 8.8 million worldwide in 2015 to more than 12 million in 2030 [3]. By 2030, the number of new cancer cases in Tanzania is projected to more than double, increasing from 37,000 cases in 2015 to over 61,000 cases[4].

Malnutrition is a nutritional condition, either acute or chronic, characterized by an excess or deficiency of nutrients, energy imbalance, and inflammatory activity. It leads to changes in body composition, impaired function, and clinical outcomes[5]. Studies indicate that the prevalence of malnutrition among cancer patients ranges from 20% to 70%, and it is estimated that 10% to 20% of cancer patient deaths worldwide are attributed to malnutrition rather than the malignancy itself[6].

Malnutrition is a major concern among cancer patients, with prevalence rates ranging from 40% to 80%. It affects approximately 15–20% of patients at diagnosis and up to 80–90% of those with advanced-stage disease. Consequently, malnourished patients tend to show lower response rates to treatment. In Latin America, studies have indicated that the prevalence of malnutrition among cancer patients ranges from 50% to 80%, whereas in Portugal, it ranges from 30% to 60%[2].

Malnutrition is more prevalent in elderly cancer patients compared to younger patients. There is a need for early integration of nutritional counselling for cancer patients, particularly for the elderly[7]. Cancer and its treatment elevate the risk of malnutrition, making early assessment of nutritional status vital for prompt nutritional intervention and improved patient survival. The disease and its therapies affect nutritional status by disrupting the metabolic system, altering taste perception, and reducing food intake. These changes can lead to physiological and psychological alterations, potentially impacting the quality of life.[8]

Tumors can alter the body’s metabolism, increasing energy expenditure while simultaneously causing anorexia, nausea, and vomiting, which reduce food intake. Chemotherapy, radiation, and surgical treatments can exacerbate these issues by causing mucositis, dysphagia, and gastrointestinal disturbances, further impairing nutritional status[9].

Despite the high prevalence of malnutrition among cancer patients, relatively few studies have explored the nutritional status of adult cancer patients[10]. The mechanisms behind cancer-related malnutrition are complex and not entirely understood. Malnutrition often arises from a combination of factors, including the increased metabolic demands of the tumor, side effects of treatment, and the patient’s diminished ability to consume and absorb nutrients. While malnutrition is recognized as a critical issue affecting cancer patients globally, leading to poor treatment outcomes and increased mortality, there is a lack of comprehensive research in Tanzania on its prevalence and associated risk factors. Existing studies have not thoroughly explored specific risk factors such as cancer stage, treatment type, and comorbidities, particularly in low-resource settings like Tanzania. This gap highlights the need for studies that not only assess the prevalence of malnutrition but also examine these critical contributors to nutritional status. Understanding these factors is crucial for designing targeted interventions to improve the health outcomes of adult cancer patients undergoing treatment.

This study aims to fill these gaps by examining the factors contributing to malnutrition among adult cancer patients undergoing treatment at Kilimanjaro Christian Medical Centre (KCMC) in Tanzania.

## Methods

### Study design

A hospital-based cross-sectional study was conducted among adult cancer patients (≥18 years) receiving treatment at the Oncology Department of KCMC Hospital from June to July 2024.

### Study area

The study was conducted in the Oncology Department at Kilimanjaro Christian Medical Centre (KCMC) Hospital, located at the foothills of Mount Kilimanjaro in Tanzania. The hospital was established in March 1971 by the Good Samaritan Foundation. It serves as a referral hospital for over 15 million people in Northern Tanzania, with a capacity of 630 official beds, 90 canvas beds, and 40 baby incubators, accommodating 500–800 inpatients. It also supports 1,852 students, 1,300 staff members, and approximately 1,000 visitors and companions. It is one of the four Zonal Consultant hospitals in Tanzania.

### Study Population and Eligibility Criteria

The study included adult cancer patients (≥18 years) receiving treatment at the Oncology Department, KCMC Hospital, who provided informed consent and had been undergoing treatment for at least six months. Patients who declined to provide consent or were critically ill were excluded from the study.

### Sample Size and Sampling Techniques

Participants were purposively selected from adult cancer patients receiving treatment at the Oncology Department, KCMC Hospital. The sample size was calculated using the Kish-Leslie formula (2015), with a 5% margin of error and a 95% confidence interval. The minimum sample size of 316 was calculated to be appropriate for this study. 323 adult cancer patients were enrolled in the study.

### Pilot Study

A pilot study was conducted with 32 adult cancer patients, representing 10% of the required sample size, to assess the feasibility, clarity, and applicability of the study tool. Four research assistants were trained on study objectives, data collection tools, informed consent procedures, participant rights, confidentiality, and interaction techniques.

### Study Variables

In this study, malnutrition was the dependent variable, influenced by several independent variables, including socio-demographic factors (age, sex, education level, marital status, residence, and occupation), clinical factors (cancer type, treatment duration, cancer stage, and treatment type), and feeding habits (food intake, mode of feeding, weight changes, and symptoms affecting food consumption). Physical activity levels were also considered to assess functional status and its impact on nutrition. The severity of malnutrition was classified using the Patient-Generated Subjective Global Assessment (PG-SGA) scoring system, with scores between 0 and 8 indicating no malnutrition and scores above 9 signifying malnutrition. This approach enables the identification of risk factors affecting nutritional status and guides targeted interventions to improve patient care, with PG-SGA providing a standardized tool to assess malnutrition severity and inform treatment decisions.

### Data collection tool and procedure

Data for this study were collected through face-to-face interviews using a structured questionnaire with a tool. The study procedures included introducing participants to the study objectives, obtaining informed consent, conducting the interviews, and providing an opportunity for participants to ask questions and clarify any doubts. Anthropometric measurements were taken following WHO guidelines using a calibrated digital scale whereby trained research assistants independently measured each participant’s weight and height, with the average of three measurements recorded. The Patient-Generated Subjective Global Assessment (PG-SGA) tool was used to assess nutritional status, with scoring based on weight history, food intake, symptoms, and physical activity levels, all evaluated according to malnutrition severity.

### Data Management and Analysis

To ensure data accuracy, data entry was double-checked, and identifiers were removed to maintain anonymity. Access to the data was restricted to authorized personnel only. Descriptive analysis was conducted by summarizing categorical variables into frequencies and percentages. For inferential analysis, Chi-square tests were performed to assess the association between malnutrition and the independent variables. Additionally, a log-binomial model was used to determine the strength of the observed associations.

## Results

### Socio-demographic characteristics of the Patients

The participants had an average age of 59.6 years and were almost evenly split between age groups, with 149 (46.1%) under 60 years and 174 (53.9%) aged 60 years or older. A slight majority were female, 169 (52.3%), while 154 (47.7%) were male. Most participants had completed primary education, 170 (52.6%), followed by those with higher education, 139 (43.3%), and a smaller proportion had informal education, 14 (4.3%). The majority were married, 218 (67.7%), while 50 (15.5%) were single and 55 (17.0%) divorced. Most participants resided in rural areas, 179 (55.4%), compared to 144 (44.6%) in urban areas. Regarding employment status, the majority were unemployed, 191 (59.1%), followed by retirees, 71 (22.0%), the employed, 51 (15.8%), and students, 10 (3.1%). In terms of health insurance, 188 (58.2%) participants had coverage, while 135 (41.8%) did not. (Table 1).

**Table 1:** Socio-demographic characteristics of the Patients (N=323).

| Variables | Total<br>N=323 |  |
| --- | --- | --- |
|  | Frequency | Percentage |
| **Age** |  |  |
| <60 | 149 | 46.1 |
| ≥ 60 | 174 | 53.9 |
| **Sex** |  |  |
| Female | 169 | 52.3 |
| Male | 154 | 47.7 |
| **Educational status** |  |  |
| Informal | 14 | 4.3 |
| Primary | 170 | 52.6 |
| Higher Education | 139 | 43.3 |
| **Marital status** |  |  |
| Single | 50 | 15.5 |
| Married | 218 | 67.7 |
| Divorced | 55 | 17.0 |
| **Residences** |  |  |
| Rural | 179 | 55.4 |
| Urban | 144 | 44.6 |
| **Occupational status** |  |  |
| Employed | 51 | 15.8 |
| Unemployed | 191 | 59.1 |
| Retired | 71 | 22.0 |
| Student | 10 | 3.1 |
| **Health Insurance** |  |  |
| Yes | 188 | 58.2 |
| No | 135 | 41.8 |

### Proportion of Malnutrition among Patients

The overall proportion of malnutrition among the patients was 234(72.5%). Specifically, 123(72.8%) of female and 111(72.1%) of male patients were malnourished, as illustrated in Figure 1. These findings underscore the critical need for targeted nutritional support and interventions to address and prevent malnutrition in this patient population.

**Figure 1:**
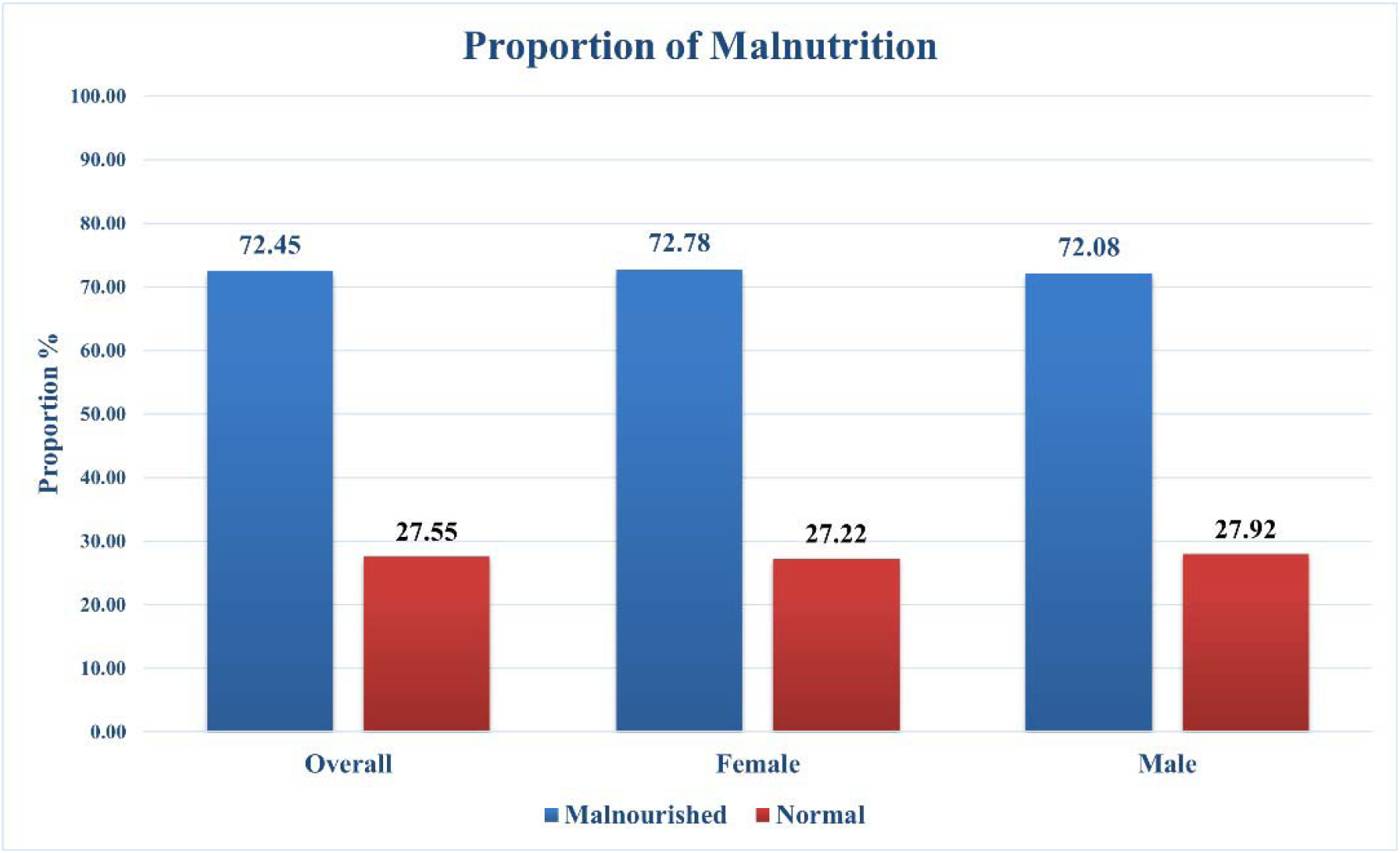
Proportion of malnutrition among adult cancer patients receiving cancer treatment at KCMC Hospital, Kilimanjaro (N=323)

### Clinical Characteristics of Patients and their Relation to Malnutrition

Table 2 details the clinical characteristics of 323 adult cancer patients, providing important insights into the prevalence and factors contributing to malnutrition among them. Among these patients, 207(74.7%) with multiple existing comorbidities were malnourished, possibly due to the additional physiological stress or dietary limitations imposed by these conditions. The lower percentage of malnourished patients in the “No comorbidity” group suggests that having multiple health conditions significantly increases the risk of malnutrition in cancer patients(p<0.05). When analyzing cancer types, 71(78.0%) of breast cancer patients were malnourished, and 61(73.5%) of those with prostate cancer were malnourished, suggesting that these cancers might involve metabolic demands or treatment side effects that adversely affect nutrition. Additionally, 38(70.4%) of colorectal cancer patients were malnourished, which is particularly significant given the direct impact of this cancer on the digestive system and nutrient absorption. Among cervical cancer patients, 15(71.4%) were malnourished, while 49(66.2%) of patients with other types of cancer also faced malnutrition, indicating a broad spectrum of cancer-related nutritional challenges.

**Table 2:**
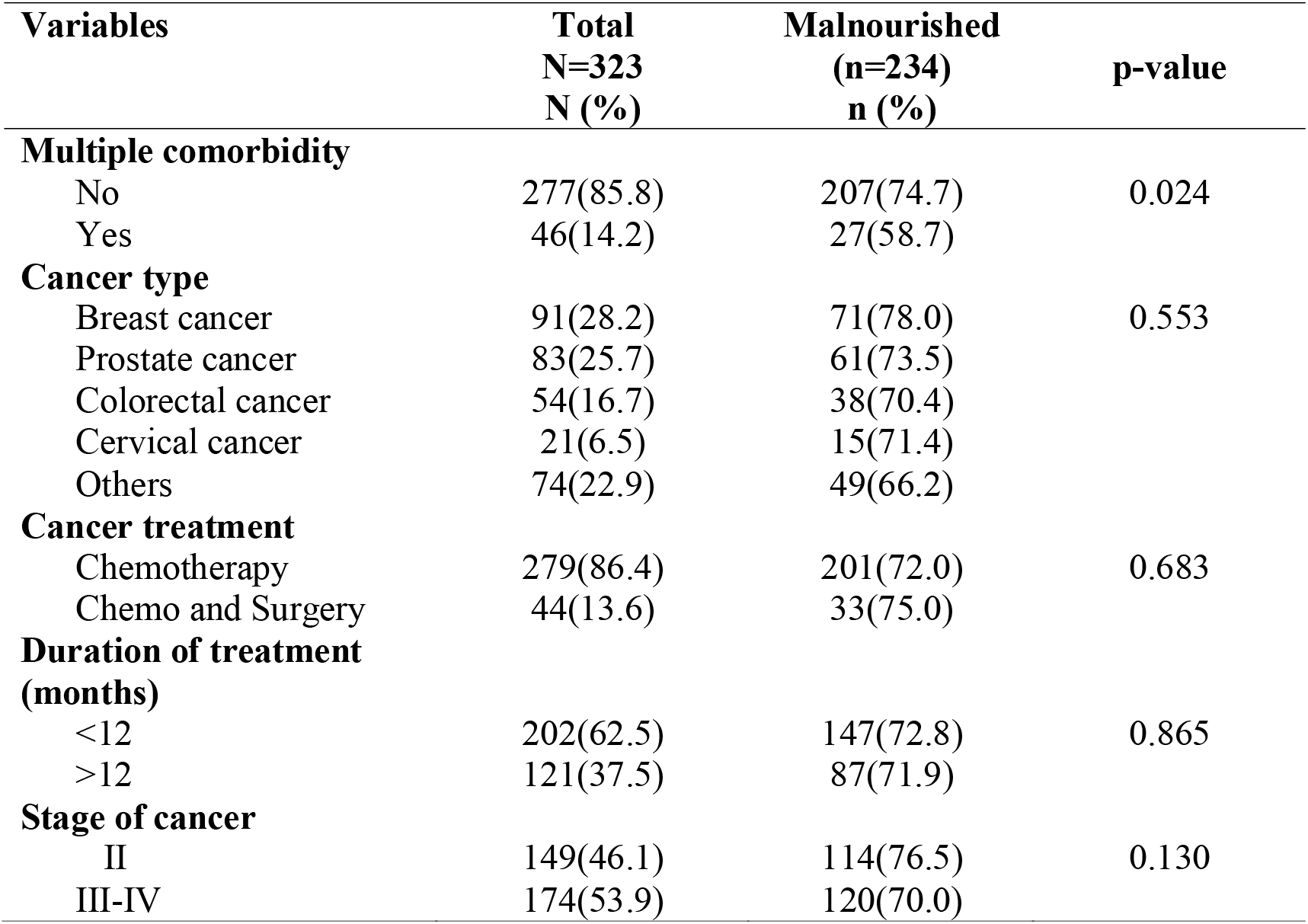
Clinical Characteristics of the Patients and their relation to Malnutrition (N=323).

Regarding cancer stages, 114(76.5%) of patients diagnosed at stage II were malnourished. This relatively even distribution suggests that malnutrition is a concern across both early and advanced stages, potentially due to both the disease itself and the effects of its treatment. In terms of treatment, a significant 33(75.0%) of the patients undergoing chemotherapy and surgery were malnourished, which often leads to side effects like nausea, vomiting, and taste changes that can reduce food intake and contribute to malnutrition.

Furthermore, 147(72.8%) of the patients who received cancer treatments within the past 12 months were malnourished. This indicates that malnutrition is prevalent both during active treatment and as a long-term consequence, underscoring the need for ongoing nutritional support and monitoring to address both immediate and persistent nutritional challenges in cancer patients.

### Feeding Habit Characteristics, activities, and functions of the Patients and their relation to Malnutrition

Of the 323 patients, 231(72.6%) reported consuming solid foods and were malnourished. This suggests that while most patients can eat a variety of foods, the quality and quantity of their diet may be insufficient. Additionally, 56(77.8%) of patients who experienced dry mouth, 108(71.5%) who had a loss of appetite, and 70(70.0%) who suffered from fatigue were malnourished. These symptoms are common side effects of cancer and its treatments, contributing to inadequate nutritional intake.

The Patient-Generated Subjective Global Assessment (PG-SGA) revealed that 142(75.5%) of patients who maintained a normal eating pattern were malnourished, and 57(75.0%) of those who had gained weight were also malnourished. Regarding physical activity,192(73.6%) of patients who could perform moderate activities were malnourished, while 17(70.8%) of those capable of normal daily functions were malnourished. Additionally, 25(65.8%) of bedridden patients were found to be malnourished.

### Factors associated with malnutrition among study participants (N=323)

This study revealed no statistically significant associations between malnutrition and examined factors. Age was not a determinant (CPR=0.9, 95% CI: 0.9-1.1, p=0.9), nor was sex (CPR=0.9, 95% CI: 0.9-1.1, p=0.9). Employment status also showed no significant influence on malnutrition, with all p-values above 0.3 (Table 4)

No significant associations were found between malnutrition and cancer-related factors. Breast, prostate, colorectal, and cervical cancers were not statistically significant predictors (p=0.1–0.6). The presence of multiple comorbidities also showed no significant relationship with malnutrition (CPR=0.8, 95% CI: 0.6-1.0, p=0.1). Similarly, cancer treatment type (CPR=1.0, 95% CI: 0.9-1.3, p=0.7) and cancer stage (CPR=0.9, 95% CI: 0.8-1.0, p=0.1) were not significantly associated with malnutrition (Table 3)

**Table 3:**
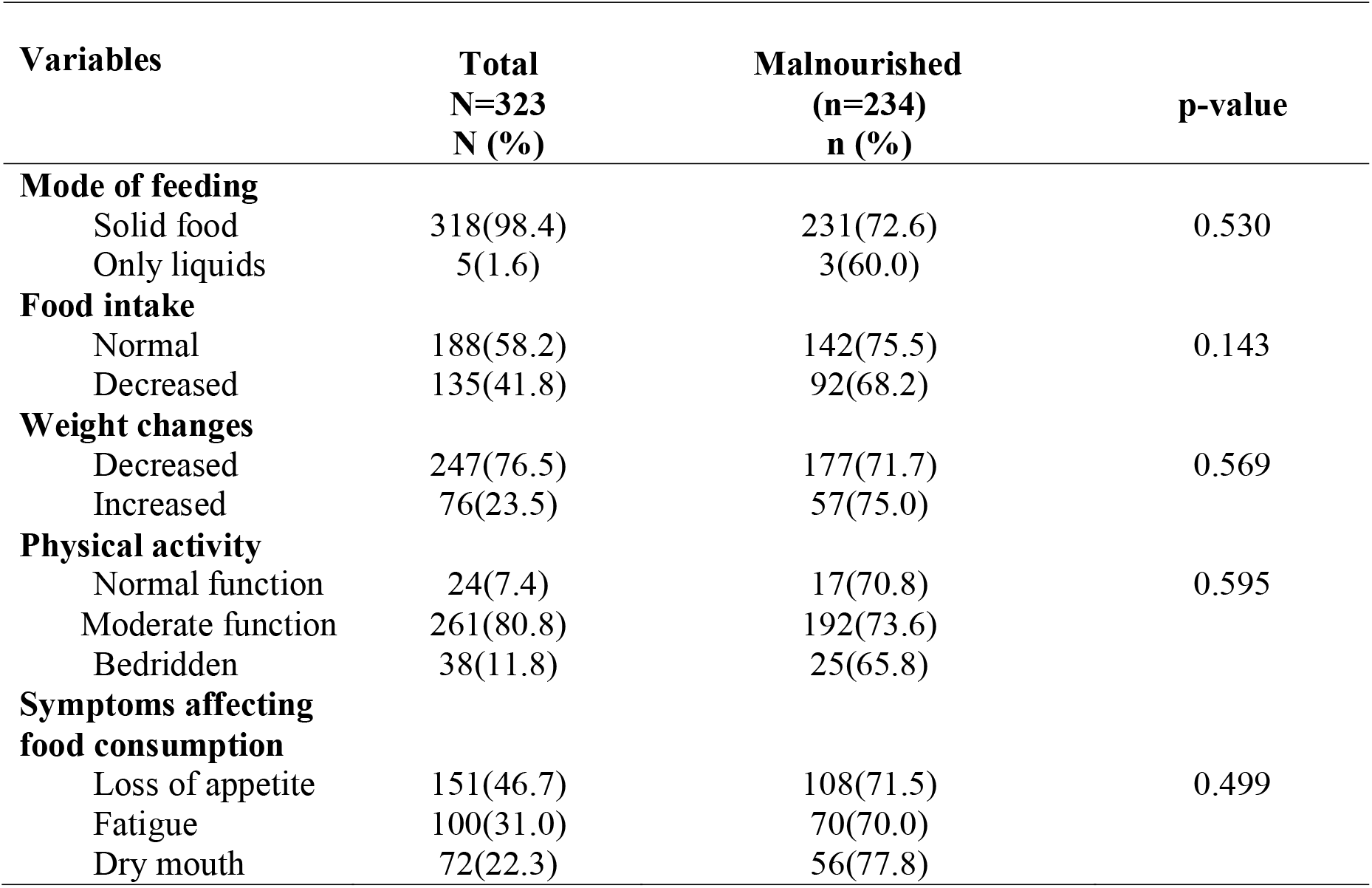
Relations between Feeding habits characteristics, activities, and functions of the Patients and their relation to Malnutrition (N=323).

Nutritional and symptomatic factors showed no significant associations with malnutrition. Decreased food intake was not significantly associated with malnutrition (CPR=0.8, 95% CI: 0.4-1.7, p=0.6). Weight changes also had no significant effect (CPR=1.0, 95% CI: 0.9-1.2, p=0.6). Among symptoms, dry mouth (CPR=1.1, 95% CI: 0.9-1.3, p=0.3), appetite loss (p=0.8), and fatigue (CPR=1.0, 95% CI: 0.8-1.2, p=0.8) showed no significant associations with malnutrition (Table 4).

**Table 4:**
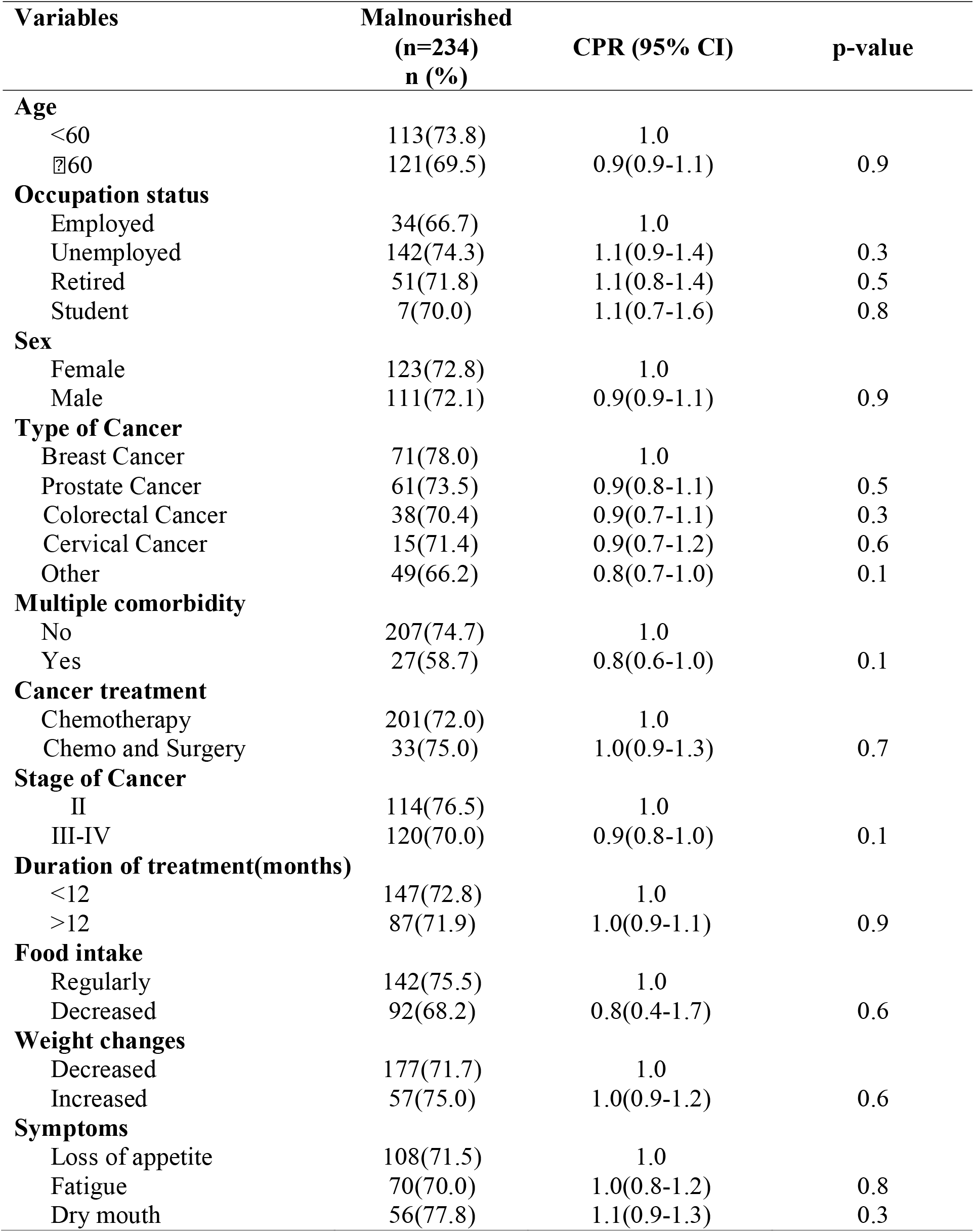
Bivariate analysis of factors associated with malnutrition.

## Discussion

The present study revealed a high prevalence of malnutrition among adult cancer patients receiving treatment at the Oncology Department of KCMC Hospital, with a proportion of 72.5%. This finding is consistent with previous studies conducted in different settings. For instance, [11] reported a malnutrition prevalence of 51.8% among cancer patients at Bugando Medical Centre, Tanzania, while [12] Found a higher rate of 83.5% in Libya. [2] Similarly, it was reported that 58.4% of adult cancer patients in Ethiopia were malnourished. The variation in these prevalence rates may be attributed to differences in socio-demographic characteristics, study populations, healthcare access, and economic conditions. These findings underscore the global burden of malnutrition among cancer patients, highlighting the urgent need for region-specific nutritional interventions to address this critical issue.

In this study, among patients who experienced weight loss, 71.7% were malnourished, and this was frequently accompanied by symptoms such as loss of appetite (71.5%), fatigue (70.0%), and dry mouth (77.8%). These results are consistent with findings from other studies. [13] Reported that 86.4% of cancer patients in Turkey experienced significant weight loss. [14] Observed that 11.1% of cancer patients in Brazil had experienced weight loss. Weight loss is a common and critical issue in cancer patients, as it is often indicative of underlying malnutrition. This underscores the importance of routine nutritional assessments and timely interventions to prevent or mitigate weight loss and its associated complications. In addition, this study found that 75.5% of patients who reported normal food intake were malnourished, while 68.2% of those with decreased food intake were also malnourished. Similar trends were observed by [2] In Ethiopia, where 28.9% of cancer patients have reduced their food intake. These findings highlight the critical role of monitoring food intake in identifying patients at risk of malnutrition. Reduced food intake is a key contributor to malnutrition, and early identification allows for prompt nutritional interventions that can improve patient outcomes, enhance recovery, and support overall quality of life. Regular assessment of dietary habits is essential in cancer care to ensure that patients receive adequate nutrition to support their treatment and recovery.

Dietary patterns also emerged as an important factor in this study. Among patients who consumed solid foods, 72.6% were malnourished, while 60% of those who consumed only liquids were malnourished. These findings align with those of [2], who found that 73.3% of cancer patients in Ethiopia consumed semisolid diets, 13.5% consumed liquid diets, and 11.1% consumed solid diets. The variation in dietary intake underscores the need for personalized nutritional interventions tailored to the specific needs of individual patients. Proper nutrition is essential for maintaining strength, improving treatment outcomes, and enhancing the overall quality of life for cancer patients. Addressing dietary intake through individualized nutritional support can help optimize treatment effectiveness and improve patient well-being.

The study also highlighted the relationship between functional impairment and malnutrition. Among patients who could perform normal functions, 70.8% were malnourished, while 73.6% of those with moderate functional ability and 65.8% of bedridden patients were malnourished. These findings are consistent with those of [15] In Northern China, where 62.4% of cancer patients experienced functional impairments, and [16] In Texas, where 88.1% of patients had impaired mobility. Functional impairments can exacerbate malnutrition by limiting a patient’s ability to prepare and consume food, thereby affecting their nutritional status. A comprehensive approach to cancer care that integrates both nutritional support and rehabilitation services is essential for addressing the multifaceted challenges faced by malnourished cancer patients. Such an approach can help improve patient outcomes, enhance recovery, and support overall quality of life.

Contrary to expectations, this study did not find significant associations between sociodemographic, clinical, and food intake characteristics with malnutrition. Several factors may explain this lack of association, including the homogeneity of the study population, which may have limited the variability in key factors such as education, income, and access to healthcare. Additionally, the cross-sectional design of the study captures only a single point in time, which may not reflect the dynamic nature of malnutrition and its potential associations over time. Longitudinal studies are needed to better understand the complex relationships between sociodemographic and clinical factors and malnutrition in cancer patients. Moreover, external factors such as healthcare infrastructure, access to nutritional supplements, and cultural influences on dietary habits may also play a role in determining nutritional status and were not fully accounted for in this study.

This study highlights the significant burden of malnutrition among adult cancer patients receiving treatment at KCMC Hospital. The prevalence of malnutrition observed in this study is consistent with findings from similar studies conducted in other regions, underscoring the global nature of the problem. The high prevalence of malnutrition among cancer patients, particularly those experiencing weight loss, reduced food intake, or functional impairments, highlights the need for targeted nutritional interventions as part of comprehensive cancer care. Early identification of malnutrition and timely implementation of personalized nutritional support can help improve treatment outcomes, support recovery, and enhance the overall quality of life for cancer patients. Future research should focus on exploring the underlying factors contributing to malnutrition in diverse populations and across different stages of cancer treatment to develop more effective interventions

### Study strength

The strengths of this study include the presence of a high response rate from the patients. The data were collected by interviewing patients, and from their medical records. The study was performed in the only oncology center in the Northern zone of Tanzania where patients are coming to this center from all over the country and from nearby countries like Kenya and Uganda. The use of the well-validated Patient-Generated Subjective Global Assessment (PG-SGA) tool for assessing malnutrition and comprehensive data collection across demographic, socioeconomic, clinical, and feeding habits features provides a young perspective on malnutrition influences. Moreover, by acknowledging the complex nature of malnutrition, the study underscores the need for a multidisciplinary approach and sets a foundation for future research to address identified gaps and limitations.

### Study limitations

The study encountered several limitations. A key issue was the scarcity of existing literature on the prevalence and risk factors of malnutrition among adult cancer patients undergoing treatment, which limited the contextual understanding of the findings. The cross-sectional design of the study prevented the establishment of causal relationships between malnutrition and its predictors. The sample was restricted to adult cancer patients at a single center, which may not fully represent all cancer patients or those receiving different types of treatments, thereby affecting the generalizability of the results. Furthermore, the study did not dig deeply into cultural and genetic factors that can significantly affect malnutrition. Given that this was a single-center study, its findings may not be broadly applicable, highlighting the need for multi-center studies to enhance generalizability. Addressing these limitations in future research will be important for gaining a more comprehensive understanding of malnutrition in cancer patients and developing effective intervention strategies.

## Conclusion

The study found that 234(72.5%) of adult cancer patients undergoing treatment were malnourished, with a higher prevalence in female patients than in males. The research emphasizes the importance of early detection of malnutrition to enable timely nutritional interventions. This highlights the critical need to include nutritionists as part of the treatment approach for adult cancer patients with a particular focus on female adult patients who exhibited a higher prevalence of malnutrition. The study also calls for further qualitative and longitudinal research to explore malnutrition in cancer patients.

## Data Availability

All data produced in the present study are available upon reasonable request to the authors

https://docs.google.com/document/d/1lBXlB2lZ5Vv00OEakDoR32sBjRi-ziwK/edit?usp=sharing&ouid=101889900679343133105&rtpof=true&sd=true

## Abbreviations

CI: Confidence Interval
CPR: Crude Prevalence Ratio
IPH: Institute of Public Health
KCMC: Kilimanjaro Christian Medical Centre
KCMUCo: Kilimanjaro Christian Medical University College
LMICs: Lower-Middle-Income Countries
PG-SGA: Patient-Generated Subjective Global Assessment
PR: Prevalence Ratio
STATA: Statistical software for data science
WHO: World Health Organization

## Declaration

- **Ethical approval**

Ethical approval for the study was obtained from Kilimanjaro Christian Medical University College (approval number PG 06/2024). Data collection permissions were obtained from the Executive Director of KCMC Hospital, the Head of the Department of Cancer, and the Oncology Department Nurse Officer in charge. Informed consent was obtained from all participants.

- **Patient consent for publication:** All participants in this study gave consent.
- **Availability of data and materials:** The dataset used is available from the corresponding author upon reasonable request
- **Competing interest:** No competing interest
- **Funding:** This was a self-funded study.

## Authors Contribution

Ephraim E. Ng’weshemi: Conceptualization of the study, Proposal writing, Data collection, Data analysis, Report writing, Manuscript writing.

Furaha Serventi: Conceptualization of the study and Supervision

Esther L. Majaliwa: Conceptualization of study and Supervision

Mary Mosha: Conceptualization of the study and Supervision across all stages of the study.

## Acknowledgement

Appreciation to KCMUCo, KCMC hospital and the Oncology Department.

## Declaration of AI assistance

We acknowledge that AI technology (ChatGPT) was partially utilized to enhance the readability and language of this work. However, the core ideas, research tasks, data analysis, findings, and interpretations remain entirely based on our original data and were independently conducted by us without any modification by AI.

